# Agent-based modeling and phylogenetic analysis suggests that COVID-19 will remain a low-severity albeit highly transmissible disease

**DOI:** 10.1101/2023.01.27.23285126

**Authors:** Juan C. Toledo-Roy, Gabriel E. García-Peña, Ana M Valdes, Alejandro Frank Hoeflich

## Abstract

The ongoing COVID-19 pandemic is still producing hundreds of thousands of cases worldwide. However, the currently dominant Omicron variant (and its sub-variants) have proven to be less virulent than previous dominant variants, resulting in proportionately fewer severe cases, hospitalizations and deaths. Nonetheless, a persistent concern is that new mutations of the SARS-CoV-2 virus may yet produce more virulent variants. In the present study we provide evidence supporting the hypothesis that this is unlikely, and that COVID-19 will remain a low-severity although highly transmissible disease. Three complementary pieces of evidence support our argument. First, empirical observations suggest that the transmission advantage that Omicron (sub)variants enjoy is in large part due to their cell tropism in the upper respiratory tract, which renders them less virulent. Second, when a negative link between transmissibility and virulence is included in agent-based epidemiological models, viruses evolve towards lower virulence. Third, genetic diversification of SARS-CoV-2 suggests that epistasis in the Omicron family reduces the diversity of successful variants. Taken together these observations point to a high likelihood that the severity of COVID-19 will remain sufficiently low for an endemic status to be reached, provided that vaccination campaigns and sensible hygiene and social measures continue worldwide, as suggested by the World Health Organization.

## Introduction

In December 2019, the first reports of individuals affected with a new serious respiratory illness in Wuhan triggered alerts and international concern. In 2020 the novel disease developed into the COVID-19 pandemic caused by the SARS-CoV-2 coronavirus [1]. Since then, the coronavirus has infected more than 600 million people, causing over 6 million deaths worldwide, as well as devastating social and economic impacts [2,3].

The original virus isolated from the first patients in Wuhan, China, has evolved and diversified into variants and subvariants of concern [4–6]. While Omicron is the most transmissible variant seen to date, at the same time it is less virulent than previous variants. Because of this, the Omicron era of dominance has resulted generally in less severe disease, and hospitalizations and deaths have not increased worldwide even if the number of reported cases is substantially higher (**Figure 1**). Indeed, quantitative studies have shown that the risk of hospitalization and/or death are substantially lower for Omicron compared to Delta [7].

**Fig. 1:**
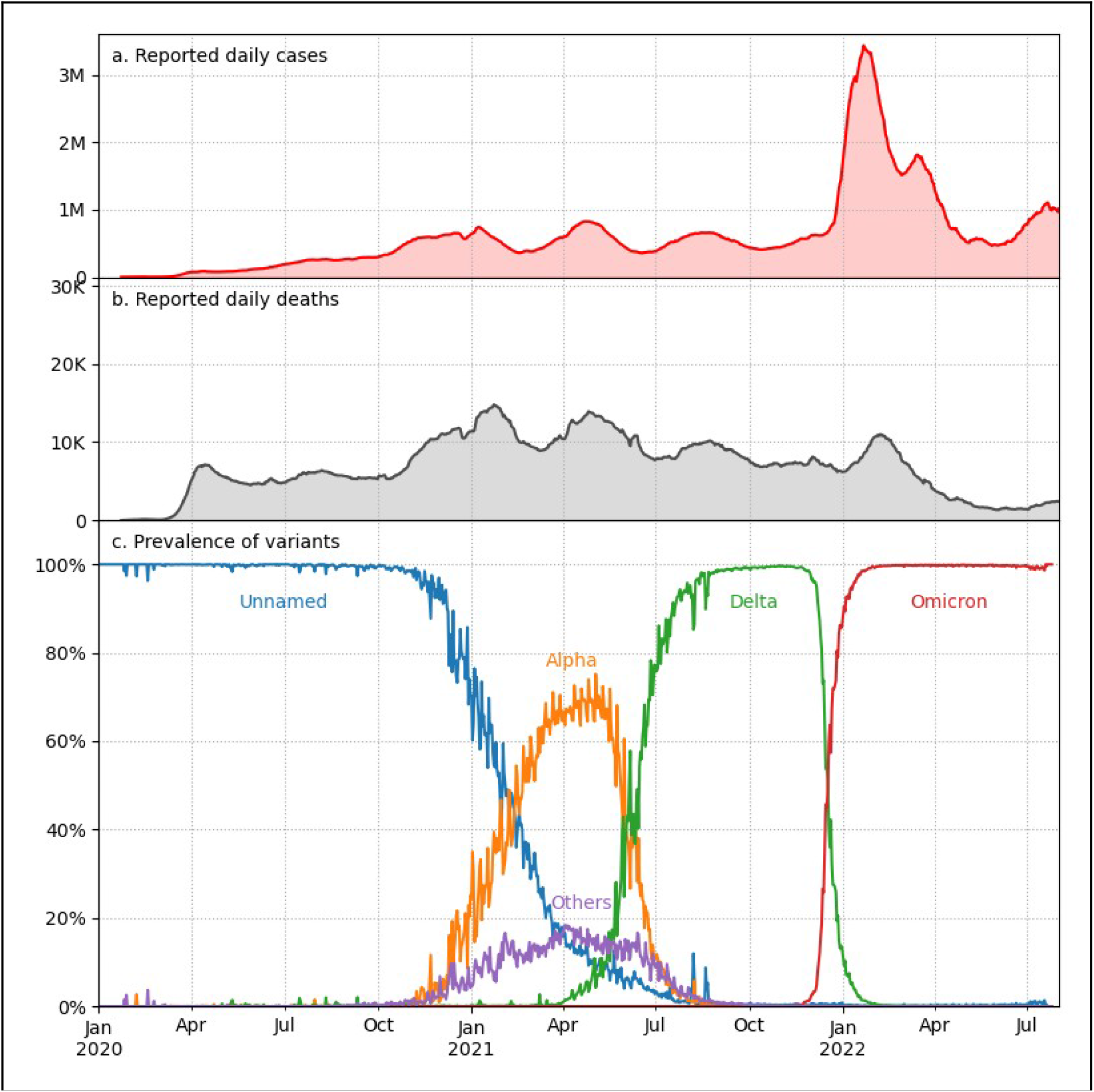
Reported cases, deaths, and prevalence of variants during the COVID-19 pandemic. **a**. Reported daily cases of COVID-19 worldwide. **b**. Reported daily deaths of COVID-19 worldwide. Cases and deaths data from Our World In Data.org (https://ourworldindata.org/). **c**. Worldwide prevalence of variants of SARS-CoV-2 (data from GISAID). “Others” includes named variants labeled as Beta, Gamma, Epsilon, Eta, Iota, Kappa, Lambda, Mu, Theta, Zeta and Gh_490r by GISAID.

While vaccination has undoubtedly contributed to the reduced severity of the disease, many studies suggest that the Omicron variant is also inherently less severe than Delta. Accounting for demographics, previous immunizations and vulnerability of the population, the hypothesis that Omicron produces milder COVID-19 than the predecessors has been corroborated by studies in South Africa [8,9], United Kingdom [10], and Denmark [11].

In this paper we argue that the next emerging variants of SARS-CoV-2 won’t be more severe than the current Omicron sub-variants even if they are more transmissible, based on three testable arguments:

1. Omicron’s tropism for the upper respiratory tract has helped it to become more easily transmissible, but this has also made it less virulent, establishing a negative link between these two properties.
2. Because of the above negative link, low-virulence variants have a fitness advantage that has allowed them to quickly become dominant, making new higher-virulence variants evolutionarily unlikely.
3. Links among the mutations of Omicron (epistasis) and selection for high transmission reduce genetic variability, which is the source for mutations that may increase virulence.

## Results

### 1. Omicron’s cellular tropism increases transmissibility and decreases virulence

The mechanisms by which Omicron has achieved this unprecedented transmissibility is also key to reducing its severity. Experimental studies on tissue cultures demonstrate that Omicron has higher tropism to infect cells from the nose and trachea (upper respiratory system) than from the lungs [12].

This shift in the cell tropism of Omicron compared to the Delta variant and its predecessors is further explained by experimental studies on SARS-CoV-2 [13] which have demonstrated that the coronavirus can enter the cells by two pathways, the (TMPRSS2) transmembrane protease serine 2 pathway and the (ACE2) Angiotensin Converting Enzyme 2 pathway. Unlike other variants of SARS-CoV-2, Omicron is inefficient in using the protease TMPRSS2 pathway, and instead infects cells by using the ACE2 pathway [14]. Many cells in the upper respiratory system (nose and trachea) have ACE2 receptors but not the TMPRSS2 enzyme, whereas in the lung parenchima ACE2 receptors are scarce and TMPRSS2 abundant [12,14].

Using the ACE2 pathway allows Omicron to enter the cells and reproduce faster than previous variants in the upper respiratory system, but also inhibits its projection into the lungs, where it has a substantially larger capacity for damage.

By being closer to the nose and mouth of infectious patients, Omicron viruses can more easily reach susceptible individuals and rapidly spread among a population. This represents an evolutionary advantage for the virus, as it can infect more people than Delta with the same amount of virus produced by the host. Indeed, the viral load of Omicron in contagious individuals has been found to not be substantially higher than the load of Delta [15,16].

Furthermore, behavioral responses to severe COVID-19, both at the individual and societal levels, act against the evolution of more virulent variants. For example, it is likely that people with less severe symptoms will spread the virus more easily than patients with severe disease isolated at home or in a hospital [17]. Similarly, social responses, including social distancing, work and public activity restrictions and lockdowns, are more intense during outbreaks of a disease perceived as severe [18].

### 2. The impact of the link between disease severity and transmissibility on the fitness landscape

The link between high transmissibility and low severity, which is a consequence of the tropism towards the upper respiratory tract, constitutes an important synergy in the evolutionary landscape of the virus. To further explore this effect we have developed a computational agent-based model (ABM).

The ABM was designed to test the impacts that disease severity and transmissibility have on the dynamics of the epidemic, and to illustrate the consequences when these properties are linked. Models of this type are essentially a stochastic and more complex version of the classical SIR model, and have often been used to explore aspects of epidemic dynamics [19,20]. The model simulates the evolution of an epidemic in a population of interacting agents with SIR-like rules, but implementing an architecture to simulate several possible features of the pathogen that can impact agent transmission and behavior. See Methods for details.

We carried out computer simulations of this ABM in order to explore how the fitness landscape depends on two characteristics of the virus: the inherent transmissibility, which accounts for the virus’ intrinsic transmission potential, including affinity to human cells and immune system escape; and the disease severity caused by the pathogen. The main result is shown in Figure 2, which shows the simulated virus fitness landscape for a collection of variants for a range of values of these two parameters.

**Fig. 2:**
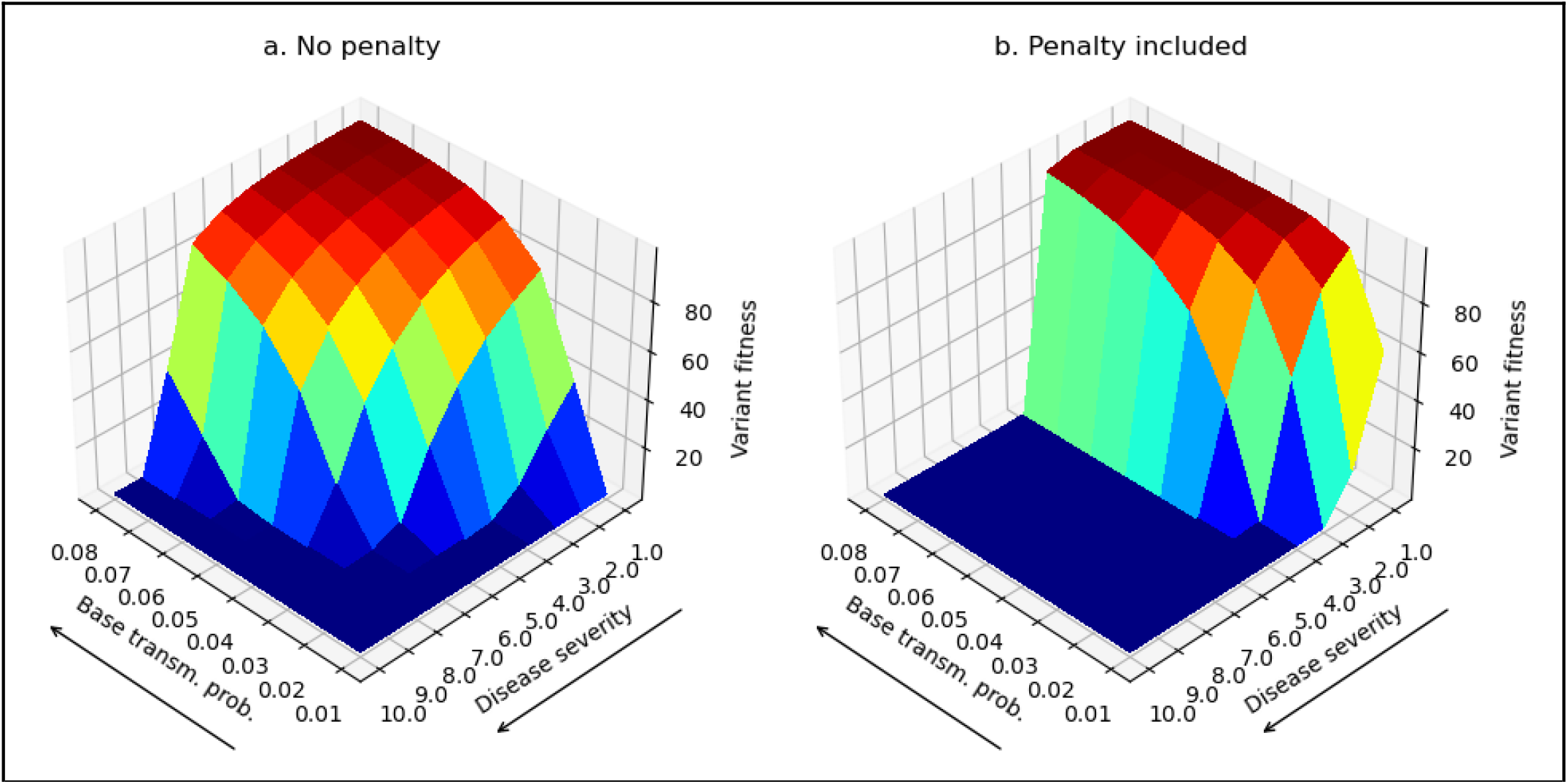
Simulated fitness landscapes resulting from the agent-based computer model. **a**. Fitness landscape when no explicit transmission penalty is associated with severity is included in the agent-based model. **b**. Landscape for the case where an explicit transmission penalty is included. In both panels, the left horizontal axis shows the base transmission probability of each variant, while the right horizontal axis corresponds to disease severity. The vertical height indicates how successful each variant is in infecting the simulated population.

The base case where no explicit transmission penalty is associated with disease severity is shown in Figure 2a. We see that the global optimum of the fitness landscape occurs at high inherent transmissibility and low severity. However, many severe variants near this optimum are also relatively competitive, and this occurs over a wide range of parameter combinations (the flat red plateau near the upper corner). Thus, variants with a higher severity can potentially remain competitive vis-a-vis others with lower severity, as long as they have a slight advantage in base transmissibility.

To explicitly model the aforementioned negative link between transmissibility and severity, we introduce a penalty to the inherent transmissibility of more virulent variants; (see *Methods* for details). When this is included in the model (see Figure 2b), the fitness landscape heavily shifts towards low severity variants, and the aforementioned wide plateau disappears.

Hence, higher severity variants (those that infect the lower respiratory system) become no longer competitive, and the landscape becomes dominated by low-severity variants, as has happened with the Omicron variant in reality. This shift in the fitness landscape makes it unlikely that a new higher severity variant can displace the current low-severity Omicron variants.

### 3. Omicrons’ epistasis reduces its phylogenetic diversity

Omicron has an outstanding number of new mutations compared with previous variants [21]. Studies on the genetic variability of SARS-CoV-2 suggest that epistasis between mutations is shaping the possibility for future mutations in the spike protein, and can potentiate the affinity of the Receptor Binding Domain to the ACE2 receptors [**22**]. For example, these authors show that mutations such as Q498R may substantially reduce ACE2 affinity, whereas it has a 25-fold enhancement affinity when present with other mutations like the N5OIY.

Therefore, the many mutations in the Omicron lineage may interact synergistically, making SARS-CoV-2 highly transmissible but less virulent, and, importantly, reducing the combinations of mutations that can be successful.

To assess this third argument, we quantified the phylogenetic diversity (PD) of Omicron, Delta and Alpha variants, observed in the global phylogeny of SARS-CoV-2 [23]. Then, we compared the PD of each variant with the number of COVID-19 cases reported in periods where each variant was dominant (with a prevalence > 50 %); (see *Methods* for further details).

Every case of COVID-19 represents an individual infected with SARS-CoV-2, and an event in which viruses reproduce and mutations accumulate [24]. Therefore, the higher the number of individuals infected with a variant the more phylogenetic diversity we expected to observe (Figure 3). The results of this quantitative exercise show that (λ) the ratio of phylogenetic diversity for each COVID-19 case is lower for Omicron (λ = 0.0091) than for Delta (λ = 0.029) and Alpha (λ = 0.012)(see methods).

**Fig. 3:**
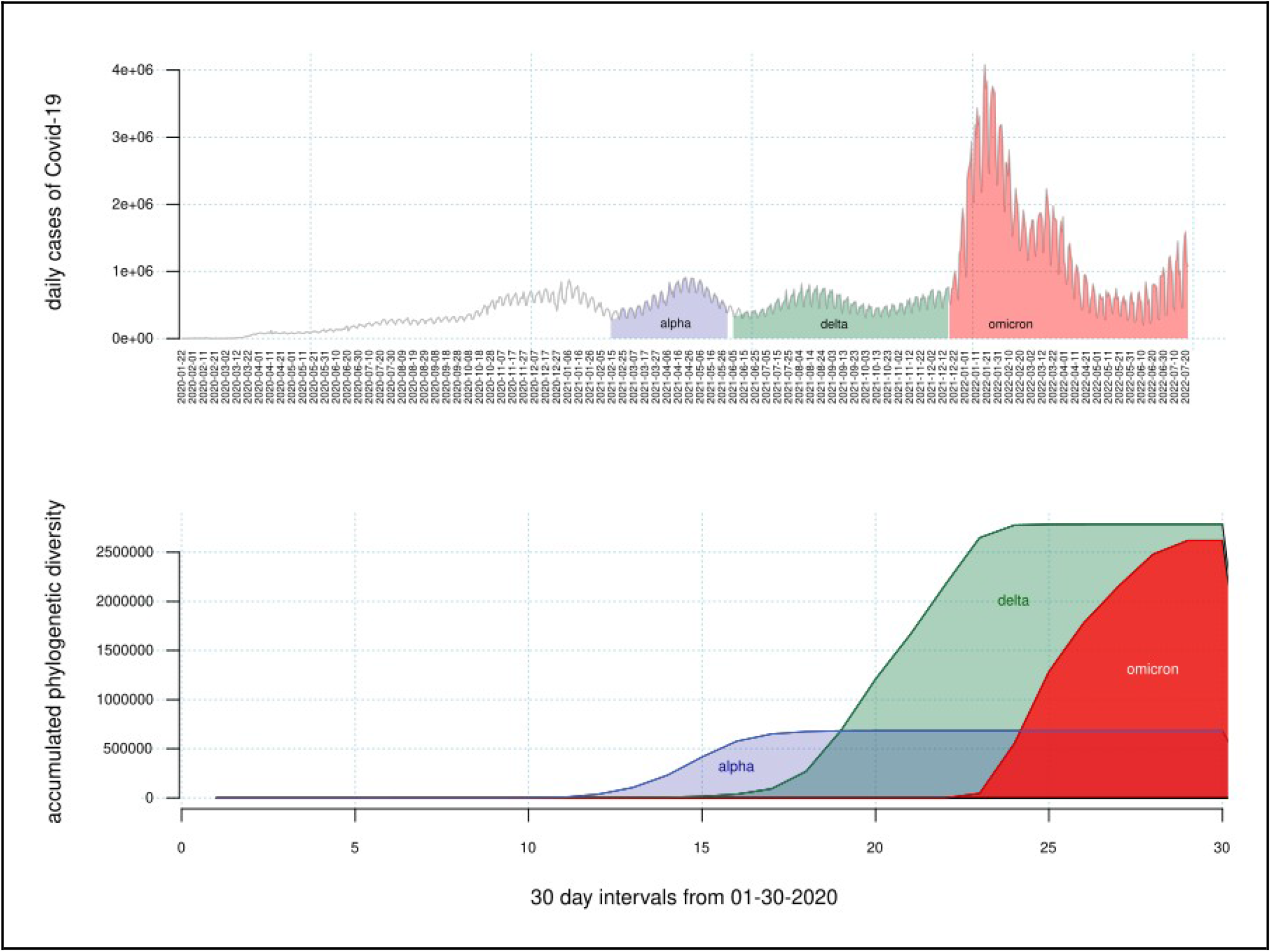
Associated cases worldwide and accumulated phylogenetic diversity of the Alpha, Delta and Omicron variants. (*Top*) Daily cases of COVID-19 across the world. Polygons highlight periods where one variant was > 50 % prevalent. During the Omicron period 294,227,135 people were infected; 97,919,266 during the period with delta, and 58,886,093 with alpha. (*Bottom*) Accumulated Faith’s phylogenetic diversity of the variants across the world. Compared to delta, Omicron has substantially less genetic variation than what is expected if each COVID-19 case is a chance for mutation.

Considering that the samples mainly capture the diversity of viruses that were successful in infecting a substantial number of individuals and the unsuccessful ones do not appear in the record of samples, the above evidence supports that Omicron is under stronger selection than were Delta and Alpha (Figure 3). When normalized for the number of infected individuals, Omicron exhibits less phylogenetic diversity, and a plausible explanation is that epistasis shapes the successful combinations of mutations in Omicron that makes the virus highly transmissible.

## Discussion

The shifted tropism of Omicron for the upper respiratory tract has helped it reach a higher transmissibility, allowing it to displace previous variants in human populations. At the same time, this shift has also resulted in a reduction in its virulence. This implies that high-transmissibility and low-severity is an evolutionary optimum towards which the virus is evolving.

By including this link, agent-based epidemiological simulations show that the fitness landscape shifts towards low-virulence, making it unlikely that higher virulence variants emerge in the future through novel mutations. Moreover, if selection favoring highly transmissible lineages is strong, as it seems to be, it is unlikely that a new more virulent variant can displace the dominant ones.

This reinforces the idea that Omicron is near an evolutionary optimum, making new more virulent variants less likely to emerge; specially if specific combinations of mutations are selected to function in synergy.

We also point out that, although COVID-19 is becoming a low-severity disease, this does not mean that we can safely ignore its presence, as it is still a very transmissible disease which can have a large social impact simply due to its ease of transmission. As pointed out by the WHO, continued and expanded vaccination campaigns as well as assessment of vaccine’s effectiveness are still crucial in reducing severe cases and deaths.

Other preventive measures such as social distancing in potentially high-transmission scenarios and good respiratory hygiene should become the new norm for our societies. This will help to minimize the collective toll that a highly transmissible disease can have, even if it is not as severe.

Finally, it is important to note that the long-term and neurological consequences of the disease are still not well understood, especially in the case of the most recent variants. In these terms, epidemiological surveillance and clinical and molecular studies on genetic variants and sub variants are fundamental tools to understand SARS-CoV-2 evolution, and be prepared for its further implications. Our quantitative analysis suggests a scientific silver lining for the end of this pandemic.

## Methods

Data on the number of COVID-19 cases and deaths worldwide were obtained from Our World in Data (https://ourworldindata.org/).

### Phylogenetic analysis

We obtained a phylogenetic tree for each of the variants, by pruning the global phylogeny of SARS-CoV-2 [23]. In a pruned tree we included the terminals labeled as variant of interest (Omicron, Delta or Alpha) by the program of variants surveillance in GISAID (https://gisaid.org), and included the reference sequence EPI_ISL_402124 as outgroup, which corresponds to an isolate from Wuhan, China in 2019.

Then we estimated phylogenetic diversity PD in each of our pruned trees, as Faith’s phylogenetic diversity, which is a conventional measure of phylogenetic diversity traditionally used for conservation studies [25]. *PD* is the sum of all the branch lengths in a phylogenetic tree.

The ratio of phylogenetic diversity by each COVID-19 case (λ) was calculated as Faiths’ phylogenetic diversity (*PD*) of the variant (*v*) divided by the number of COVID-19 cases accumulated when the variant was > 50 % prevalent (∑ *c*_*v*_), per equation (1):

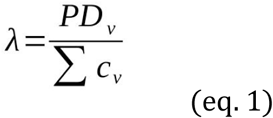

Phylogenetic analyses were performed using the R package APE [**26**].

### Agent-Based Modeling

We used an agent-based model to probe the impact of the link between transmissibility and severity on the fitness landscape. We carried out computer simulations of the spread of a contagious disease in a population of N = 10,000,000 simulated agents. Each simulated virus variant is defined by two parameters: its base transmission probability per encounter (base_transm_prob, similar to the β parameter in the SIR model) which is chosen in the range 4% to 12%, and its severity (disease_severity) which is defined as a numeric value between 1 (low severity) and 10 (high severity).

All simulated variants share the following additional parameters: an incubation period (time between infection and symptoms onset) of 3 days, a latency period of (time between infection and onset of infectiousness) of 2 days, which makes some pre-symptomatic contagion possible, a duration of infectiousness of 6 days, and a 1% infection fatality rate.

At the start of the simulation, a small number of agents (1,000) are randomly selected and infected with the variant being simulated (one simulation is run for each variant). Then, on every iteration, every agent has between 0 and base_encounters = 5 random encounters with another agent during which transmission may occur. The actual number depends on the diseases’s severity as a decreasing quadratic function, equation (2):

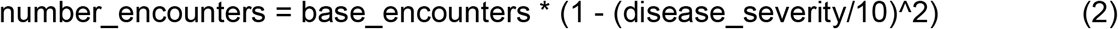

For each encounter where one agent is infected and contagious and the other is susceptible, the latter is infected with a probability given by equation (3):

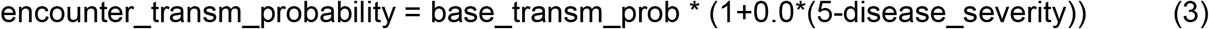

Hence, disease severity is penalized if it is above 5, and boosted if it is below 5. This codifies the link between transmissibility and severity into the simulated epidemic dynamics.

As time evolves in discrete one-day iterations, an infected agent follows the disease progression outlined by the various periods: they become contagious 2 days after infection, then symptomatic 3 days after infection (1 day after becoming contagious), and remain symptomatic for 6 more days. On each day there is a small probability (the daily fatality rate) that they die from the disease; in that case the agent is removed from the simulation. Once the disease has run its course the agent stops being contagious and is no longer susceptible to infection and is thus no longer relevant to the epidemic dynamic (we do not simulate re-infection to prevent recurrent waves).

For each variant in the explored parameter space, we simulate a new, full epidemic until no infectious agent is left (all agents are either cured of the disease, still susceptible, or diseased). Then, the variant’s success is measured by counting how many agents in the population ever became infected: the higher this number, the more successful the variant, which we interpret as a measure of its fitness in the fitness landscapes shown.

## Data Availability

All data produced in the present study are available upon reasonable request to the authors

## Data availability

Analyzed data is available publicly from the cited sources, or can be obtained upon request to the corresponding authors.

## Code availability

The C++ code implementation of the agent-based model developed for this study is available publicly at https://github.com/meithan/epidemic.

## Acknowledgements

JCTR acknowledges financial support from project PAPIIT-IA103121.

## Author contributions

All authors contributed equally to the design and development of the study as well as to writing the manuscript. JCTR developed the agent-based model and carried out the simulations. GEGP carried out the phylogenetic analysis.

## Materials & Correspondence

code & data requests may be addressed to JCTR or GEGP

